# Clinical evaluation of an immunochromatographic IgM/IgG antibody assay and chest computed tomography for the diagnosis of COVID-19

**DOI:** 10.1101/2020.04.22.20075564

**Authors:** Kazuo Imai, Sakiko Tabata, Mayu Ikeda, Sakiko Noguchi, Yutaro Kitagawa, Masaru Matsuoka, Kazuyasu Miyoshi, Norihito Tarumoto, Jun Sakai, Toshimitsu Ito, Shigefumi Maesaki, Kaku Tamura, Takuya Maeda

**Affiliations:** COVID-19 Task Force, Self-Defense Forces Central Hospital, Tokyo, Japan; Department of Infectious Disease and Infection Control, Saitama Medical University, Saitama, Japan; Center for Clinical Infectious Diseases and Research, Saitama Medical University, Saitama, Japan; Department of Clinical Laboratory, Saitama Medical University, Saitama, Japan

## Abstract

**Background:** We evaluated the clinical performance of an immunochromatographic (IC) IgM/IgG antibody assay for severe acute respiratory syndrome coronavirus 2 (SARS-CoV2) and chest computed tomography (CT) for the diagnosis of Coronavirus disease 2019 (COVID-19).

**Methods:** We examined 139 serum specimens collected from 112 patients with COVID-19 and 48 serum specimens collected from 48 non-COVID-19 patients. The presence of IgM/IgG antibody for SARS-CoV2 was determined using the One Step Novel Coronavirus (COVID-19) IgM/IgG Antibody Test. Chest CT was performed in COVID-19 patients on admission.

**Findings:** Of the139 COVID-19 serum specimens, IgM was detected in 27.8%, 48.0%, and 95.8% of the specimens collected within 1 week, 1–2 weeks, and >2 weeks after symptom onset and IgG was detected in 3.3%, 8.0%, and 62.5%, respectively. Among the 48 non-COVID-19 serum specimens, 1 generated a false-positive result for IgM. Thirty-eight of the 112 COVID-19 patients were asymptomatic, of whom 15 were positive for IgM, and 74 were symptomatic, of whom 22 were positive for IgM and 7 were positive for IgG. The diagnostic sensitivity of CT scan alone and in combination with the IC assay was 57.9 % (22/38) and 68.4% (26/38) for the asymptomatic patients and 74.3% (55/74) and 82.4% (61/74) for the symptomatic patients, respectively.

**Conclusion:** The IC assay had low sensitivity during the early phase of infection, and thus IC assay alone is not recommended for initial diagnostic testing for COVID-19. If RT-qPCR is not available, the combination of chest CT and IC assay may be useful for diagnosing COVID-19.

## Introduction

The severe acute respiratory syndrome coronavirus 2 (SARS-CoV2) epidemic, which causes the novel coronavirus disease 2019 (COVID-19), was first reported in December 2019 in Wuhan, China (1), and it has since been declared a pandemic by the World Health Organization. The ongoing outbreak is a global threat to human health. Quantitative reverse-transcription polymerase chain reaction (RT-qPCR) analysis for SARS-CoV2 is considered the gold standard for diagnosing COVID-19. RT-qPCR has been used to analyze specimens from the upper and lower respiratory tracts for clinical diagnosis during outbreaks of other diseases, but it has not been performed widely in the clinical setting because it requires special equipment, a time-consuming protocol, and highly skilled laboratory technicians. In addition, because RT-qPCR requires samples from the upper and lower respiratory tracts, the process of collecting samples and extracting RNA increases the risk of exposure to viral droplets. Therefore, an alternative diagnostic test to RT-qPCR is desirable for the clinical management of COVID-19.

In studies conducted in China, chest computed tomography (CT) scans were widely utilized as a diagnostic tool for COVID-19 (2-4). Lung involvement can be detected in patients with COVID-19 on a CT scan in advance of the symptoms typical for pneumonia (5) and a positive result on RT-qPCR (6, 7). The common radiological characteristics of COVID-19 pneumonia on chest CT have a diagnostic sensitivity of 73%–93% and a specificity of 24%–100% in distinguishing COVID-19 from other forms of viral pneumonia (2, 8). An immunochromatographic (IC) assay for IgM and IgG antibodies against the virus is widely accepted as a point-of-care test because it is an easy-to-perform, rapid, and high-throughput method for diagnosing viral infections. Recently, several commercial IC assays that detect IgM/IgG antibodies against SARS-CoV2 have become available for use in the clinical setting. However, their clinical usefulness has yet to be thoroughly evaluated. Here, we describe the clinical performance of an IC assay in comparison with that of chest CT.

## Materials and Methods

### Patients with COVID-19 and their clinical specimens

Patients with laboratory-confirmed COVID-19 who were referred to the Self-Defense Forces Central Hospital and Saitama Medical University Hospital in Japan from February 11 to March 31, 2020 were enrolled in this study. All patients were examined by RT-qPCR for SARS-CoV2 using pharyngeal and nasopharyngeal swabs collected at public health institutes or hospitals in accordance with the nationally recommended method in Japan (9). Chest CT was performed on the day of admission. Serum specimens were collected on the day of admission and during hospitalization. Clinical information was collected from the medical records. The CT findings were evaluated by a radiologist to determine the specific features caused by COVID-19 (2).

### Negative samples from patients with non-COVID-19

To evaluate the analytical specificity of the IC assay, we used serum samples collected from patients at Saitama Medical University Hospital, Japan, from April to October 2019, before SARS-CoV2 was first reported in China. Clinical information was collected from the medical records, and all serum samples were stored at −80°C before use in the IC assay.

### Definition

Asymptomatic cases were defined as patients with no history of clinical signs or symptoms. Symptomatic cases were defined as patients showing the clinical symptoms of COVID-19: fever, cough, nasal discharge, diarrhea, malaise, dyspnea, tachypnea, peripheral capillary oxygen saturation <93%, and need for oxygen therapy. The day of onset was defined as the first day of symptoms caused by COVID-19 in the symptomatic patients or the day of the first positive RT-qPCR result for upper respiratory specimens in the asymptomatic patients.

### Detection of IgM and IgG antibodies for SARS-CoV2

IgM/IgG antibody tests for SARS-CoV2 were performed using the One Step Novel Coronavirus (COVID-19) IgM/IgG Antibody Test (Artron, Burnaby, Canada) according to the manufacturer’s instructions. In brief, 10 μL serum was added to the sample port of the IC assay and was incubated for 20–30 s. Subsequently, 2 drops of sample buffer were added to the same sample port, and the results were interpreted after a 15–20 min incubation. The presence of only the control line indicates a negative result; the presence of both the control line and the IgM or IgG antibody line indicates a positive result for IgM or IgG antibody, respectively.

### Ethical statement

The study design and protocol were reviewed and approved by the Institutional Review Board of the Japan Self-Defense Forces Central Hospital (Approval No. 01-011) and Saitama Medical University Hospital (Approval Nos. 19136 and 20001).

## Results

### Sensitivity and specificity of the IC assay for COVID-19

IgM and IgG antibodies for SARS-CoV2 could be detected by the IC assay. In total, 139 serum samples were collected from 112 patients with laboratory-confirmed COVID-19 and were used as positive controls for the IC assay in this study. The medium period from onset to serum collection was 6 days (interquartile range [IQR], 3–13 days). The results of the IC assay for serum specimens are shown in Table 1. The serum samples were subdivided into three groups according to sample collection times: within 1 week (n = 90), 1–2 weeks (n = 25), and >2 weeks after onset (n = 24). IgM antibody was detected in 60 (43.2%) of the 139 serum samples collected and IgG antibody was detected in 20 (14.4%) specimens. All IgG antibody-positive samples were also positive for IgM antibody in the IC assay. Thus, the sensitivity of the IC assay was calculated to be 43.2% for all serum specimens. IgM antibody was detected in 27.8% (25/90) of specimens collected within 1 week of onset, 48.0% (12/25) collected within 1–2 weeks, and 95.8% (23/24) collected >2 weeks after onset. The corresponding detection rates for IgG antibody were 3.3% (3/90 specimens), 8.0% (2/25 specimens), and 62.5% (15/25 specimens).

**Table 1:** IC assay for IgM and IgG antibodies using COVID-19–positive serum specimens

|  | Total<br>N = 139 | Time from symptom onset to specimen collection |  |  |
| --- | --- | --- | --- | --- |
|  |  | <1 week<br>n = 90 | 1–2 weeks<br>n = 25 | >2 weeks<br>n = 24 |
| IgM | 60 (43.2%) | 25 (27.8%) | 12 (48.0%) | 23 (95.8%) |
| IgG | 20 (14.4%) | 3 (3.3%) | 2 (8.0%) | 15 (62.5%) |
| IgM + IgG | 60 (43.2%) | 25 (27.8%) | 12 (48.0%) | 23 (95.8%) |

Data are n (%).

In IC assays of the 48 non-COVID-19 serum specimens collected before the emergence of SARS-CoV2 infection, 1 specimen from a patient with Sjogren’s syndrome and rheumatoid arthritis showed a false-positive result for IgM antibody. Thus, the specificity of the IC assay was calculated to be 98.0%.

### IC assay and chest CT for patients with asymptomatic and symptomatic COVID-19

Clinical background of the 112 patients hospitalized due to COVID-19 in this study are shown in Table 2. Thirty-eight (33.9%) patients who had no COVID-19 symptoms were classified as asymptomatic and the remaining 74 (66.1%) were classified as symptomatic. Briefly, patients were aged 20–93 years (median, 67 years; IQR, 45–74 years), and 64 (57.1%) were men. All asymptomatic patients were diagnosed with COVID-19 by RT-qPCR while under quarantine in Japan. Of the 38 asymptomatic patients, median time from the first RT-qPCR–positive day to admission was 5 days (IQR, 3–6 days). Of the 74 symptomatic patients, median time from onset to admission was 5 days (IQR, 2–7 days).

**Table 2:** Clinical characteristics of patients with COVID-19 on admission

| Characteristics | Total<br>N = 112 | Asymptomatic<br>n = 38 | Symptomatic<br>n = 74 |
| --- | --- | --- | --- |
| Age (years) | 67 (45–74) | 68 (61.5–73.75) | 65 (40–74.5) |
| Sex (male) | 64 (57.1%) | 16 (42.1%) | 48 (64.8%) |
| Time from onset to admission (days) | 5 (2–7) | NA | 5 (2–7) |
| Time from first RT-qPCR-positive day to admission (days) | 3 (2–6) | 5 (3–6) | 3 (1.75–5) |

Data are n (%), or median (IQR), unless otherwise specified. NA, not applicable.

Table 3 shows the results of the IC assay and chest CT for the patients on admission. When using serum samples taken from the 38 asymptomatic patients, IgM antibody was detected in 15 (39.5%) patients on admission, and none of the patients were positive for IgG antibody. Chest CT showed abnormal lung findings consistent with the radiographic features of COVID-19 in 22 (57.9%) asymptomatic patients on admission. When the combination of IC assay and chest CT findings was used for diagnosis in the asymptomatic patients, the sensitivity was 68.4%.

**Table 3:** IC assay and chest CT findings for patients with COVID-19 on admission

|  | Asymptomatic |  |  | Symptomatic |  |  |  |
| --- | --- | --- | --- | --- | --- | --- | --- |
|  | Total<br>n = 38 | Time from first<br>RT-qPCR–positive day to<br>admission |  | Total<br>n = 74 | Time from onset to admission |  |  |
|  |  | <1 week<br>n = 35 | 1–2 weeks<br>n = 3 |  | <1 week<br>n = 53 | 1–2 weeks<br>n = 12 | >2 weeks<br>n = 9 |
| IgM | 15 (39.5%) | 14 (40.0%) | 1 (33.3%) | 22 (29.7%) | 9 (17.0%) | 4 (33.3%) | 9 (100.0%) |
| IgG | 0 (0%) | 0 (0%) | 0 (0%) | 7 (9.5%) | 2 (3.8%) | 1 (8.3%) | 4 (44.4%) |
| IgM + IgG | 15 (39.5%) | 14 (40.0%) | 1 (33.3%) | 22 (29.7%) | 9 (17.0%) | 4 (33.3%) | 9 (100.0%) |
| CT scan | 22 (57.9%) | 19 (54.2%) | 3 (100.0%) | 55 (74.3%) | 39 (73.6%) | 8 (66.7%) | 8 (88.9%) |
| IgM + IgG +<br>CT scan | 26 (68.4%) | 23 (65.7%) | 3 (100.0%) | 61 (82.4%) | 43 (81.1%) | 9 (75.0%) | 9 (100.0%) |

Data are n (%).

Of the 74 symptomatic patients, IgM antibody was detected in 22 (29.7%) patients and IgG antibody in 7 (9.5%) patients. All IgG antibody-positive patients were also positive for IgM antibody. The sensitivity of the IC assay was 17.0% (9/53) within 1 week, 33.3% (4/12) within 1–2 weeks, and 100.0% (9/9) within >2 weeks after onset. Of the 74 symptomatic patients, chest CT detected the radiographical patterns of COVID-19 in 55 (74.3%) patients on admission. The corresponding sensitivity of chest CT was 73.3% (39/53 patients), 66.7% (8/12), and 88.9% (8/9). When the combination of IC assay and chest CT was used for diagnosis in symptomatic COVID-19 patients, the corresponding sensitivity was 81.1% (43/53 patients), 75.0% (9/12), and 100% (9/9).

## Discussion

Here, we presented the analytical results of a commercial IC assay and findings of chest CT scans for patients with COVID-19. Although the IC assay showed high sensitivity for samples collected >2 weeks after symptom onset, it was less sensitive for patients who developed symptomatic COVID-19 within 1 week. Chest CT showed higher sensitivity than the IC assay for the diagnosis of COVID-19, but it did not show the specific radiological features of COVID-19 in 18.3% of symptomatic patients. Nevertheless, the combination of IC assay and chest CT slightly increased the diagnostic sensitivity for COVID-19.

Based on previous enzyme-linked immunosorbent assay (ELISA) results for IgM and IgG antibodies, only 38.3% of patients were positive for IgM antibody within the first week after onset. The detection of IgM and IgG antibodies increased rapidly from day 15 after onset (IgM = 94.3% and IgG = 79.8%) (10). Our IC assay results support these previous findings that seroconversion mainly occurred >2 weeks after onset (10). The clinical usefulness of serological tests for COVID-19 remains controversial due to the time lag between the onset of symptoms and the appearance of IgM and IgG antibodies in serum. In China and the United States, the sensitivity and specificity of serological tests for samples initially collected from hospitalized patients were 38.3%–85.4% and 100% (10-12) for ELISA and 18.4%–88.7% and 90.6%–91.7% for IC assay (13, 14), respectively.

This contradiction probably reflects differences in the timing of sampling because the clinical setting varies in each country. In the clinical setting, patients are usually diagnosed with COVID-19 within 2 weeks because they develop dyspnea and pneumonia at a median of 8 days (IQR, 5.0–13.0 days) after symptom onset (1). In this study, the median time from onset to hospitalization was 5 days (IQR, 2–7 days), which is shorter than in previous studies (median 7–15 days) (10-14). Additionally, only 29.7% of patients were diagnosed using IC assay alone, supporting the Cassaniti et al.’s conclusion that the sensitivity of IC assays remains insufficient for their use as a clinical diagnostic tool (14). Therefore, unfortunately, the IC assay alone cannot replace RT-qPCR as an acute diagnostic protocol for COVID-19, at least in the clinical setting in Japan. However, the IC assay can be used for epidemiological studies of the seroprevalence of IgM and IgG antibodies against SARS-CoV2.

Previous studies have shown that the sensitivity of CT among symptomatic patients was high (73%–97%), although specificity differed widely (24%–100%) (7, 8, 15, 16). The clinical performance of CT may vary according to differences in patient populations, disease severity, and accessibility to chest CT scans in each country. In the present study, chest CT showed higher sensitivity than the IC assay, but sensitivity was only 73.3% among the symptomatic patients who tested positive for SARS-CoV2 according to RT-qPCR. Bernheim et al. reported that the sensitivity of chest CT was low (44%) in the acute phase (0–2 days after onset) but high (91%) in the intermediate phase (3–5 days) (16). The low sensitivity of chest CT may reflect the short period of time between symptom onset to hospitalization in the symptomatic patients examined in this study. The diagnostic sensitivity was improved by combining the IC assay and chest CT (81.3%). In the present study, we did not evaluate the specificity of chest CT, but taking the high specificity of the IC assay into consideration, combining the IC assay and chest CT was considered to improve the diagnostic specificity as well. When RT-qPCR is not available or practical, the combination may be useful for diagnosing COVID-19.

The identification of asymptomatic patients with COVID-19 is important to prevent nosocomial infection. The average incubation period of COVID-19 is 5.2 days (17) but ranges from 0 to 24 days (15). It has also been reported that patients hospitalized with other diseases who did not show respiratory symptoms developed symptomatic COVID-19 and they spread SARS-CoV2 to other patients and medical workers (18). Also, the transmission of SARS-CoV2 from patients without respiratory symptoms has been reported in several countries (19-21). In the present study, chest CT showed higher sensitivity than the IC assay (57.9% vs. 39.5%, respectively), but it is not practical to perform chest CT for all hospitalized patients because of radiation exposure risk and limited medical resources (22). Although the IC assay alone may not be useful as a screening test for asymptomatic COVID-19 due to its low sensitivity, it may contribute to the prevention of nosocomial infection.

A major limitation of this study was the low number of patients. In addition, only one commercial IC kit was evaluated. The commercial IC assay verified to have the best performance in the clinical setting should be chosen for further studies. Multicenter, multi-national, prospective studies are warranted to determine the usefulness of IC assays and chest CT for diagnosing COVID-19.

## Conclusion

The sensitivity of the IC assay was low during the early phase in asymptomatic and symptomatic patients. Therefore, IC assay alone is not recommended for initial diagnostic testing for COVID-19. When RT-qPCR cannot be used, the combination of chest CT and IC assay may be useful for diagnosing COVID-19.

## Data Availability

The raw data used in this study is available from the corresponding author upon reasonable request.

## Acknowledgments

We thank everyone involved in the COVID-19 Task Force at the Self-Defense Forces Central Hospital in Japan and members who were assembled from other institutes of the Japan Self-Defense Forces.

